# Sensitivity of wastewater-based epidemiology for detection of SARS-CoV-2 RNA in a low prevalence setting

**DOI:** 10.1101/2021.08.24.21258577

**Authors:** Joanne Hewitt, Sam Trowsdale, Bridget Armstrong, Joanne R. Chapman, Kirsten Carter, Dawn Croucher, Cassandra Billiau, Rosemary Sim, Brent J. Gilpin

## Abstract

To assist public health responses to COVID-19, wastewater-based epidemiology (WBE) is being utilised internationally to monitor SARS-CoV-2 infections at the community level. However, questions remain regarding the sensitivity of WBE and its use in low prevalence settings. In this study, we estimated the total number of COVID-19 cases required for detection of SARS-CoV-2 RNA in wastewater. To do this, we leveraged a unique situation where, over a 4-month period, all symptomatic and asymptomatic cases, in a population of approximately 120,000, were precisely known and mainly located in a single managed isolation and quarantine facility (MIQF) building. From 9 July to 6 November 2020, 24-hr composite wastewater samples (*n* = 113) were collected daily from the sewer outside the MIQF, and from the municipal wastewater treatment plant (WWTP) located 5 km downstream. New daily COVID-19 cases at the MIQF ranged from 0 to 17, and for most of the study period there were no cases outside the MIQF identified. SARS-CoV-2 RNA was detected in 54.0% (61/113) at the WWTP, compared to 95.6% (108/113) at the MIQF. We used logistic regression to estimate the shedding of SARS-CoV-2 RNA into wastewater based on four infectious shedding models. With a total of 5 and 10 COVID-19 infectious cases per 100,000 population (0.005 % and 0.01% prevalence) the predicated probability of SARS-CoV-2 RNA detection at the WWTP was estimated to be 28 and 41%, respectively. When a more realistic proportional shedding model was used, this increased to 58% and 87% for 5 and 10 cases, respectively. In other words, when 10 individuals were actively shedding SARS-CoV-2 RNA in a catchment of 100,000 individuals, there was a high likelihood of detecting viral RNA in wastewater. SARS-CoV-2 RNA detections at the WWTP were associated with increasing COVID-19 cases. Our results show that WBE provides a reliable and sensitive platform for detecting infections at the community scale, even when case prevalence is low, and can be of use as an early warning system for community outbreaks.

**Highlights:**

- Over 4 months, all 0-17 new daily cases in one quarantine building, catchment 120,000 population
- Wastewater tested daily at quarantine building and downstream wastewater treatment plant, WWTP
- SARS-CoV-2 RNA detected in 95.6% (108/113) at the MIQF and 54.0% (61/113) at the WWTP
- SARS-CoV-2 RNA detections at the WWTP associated with increasing COVID-19 cases
- Probability of SARS-CoV-2 RNA detection of 87% with 0.01% total case prevalence

**Graphical Abstract:** 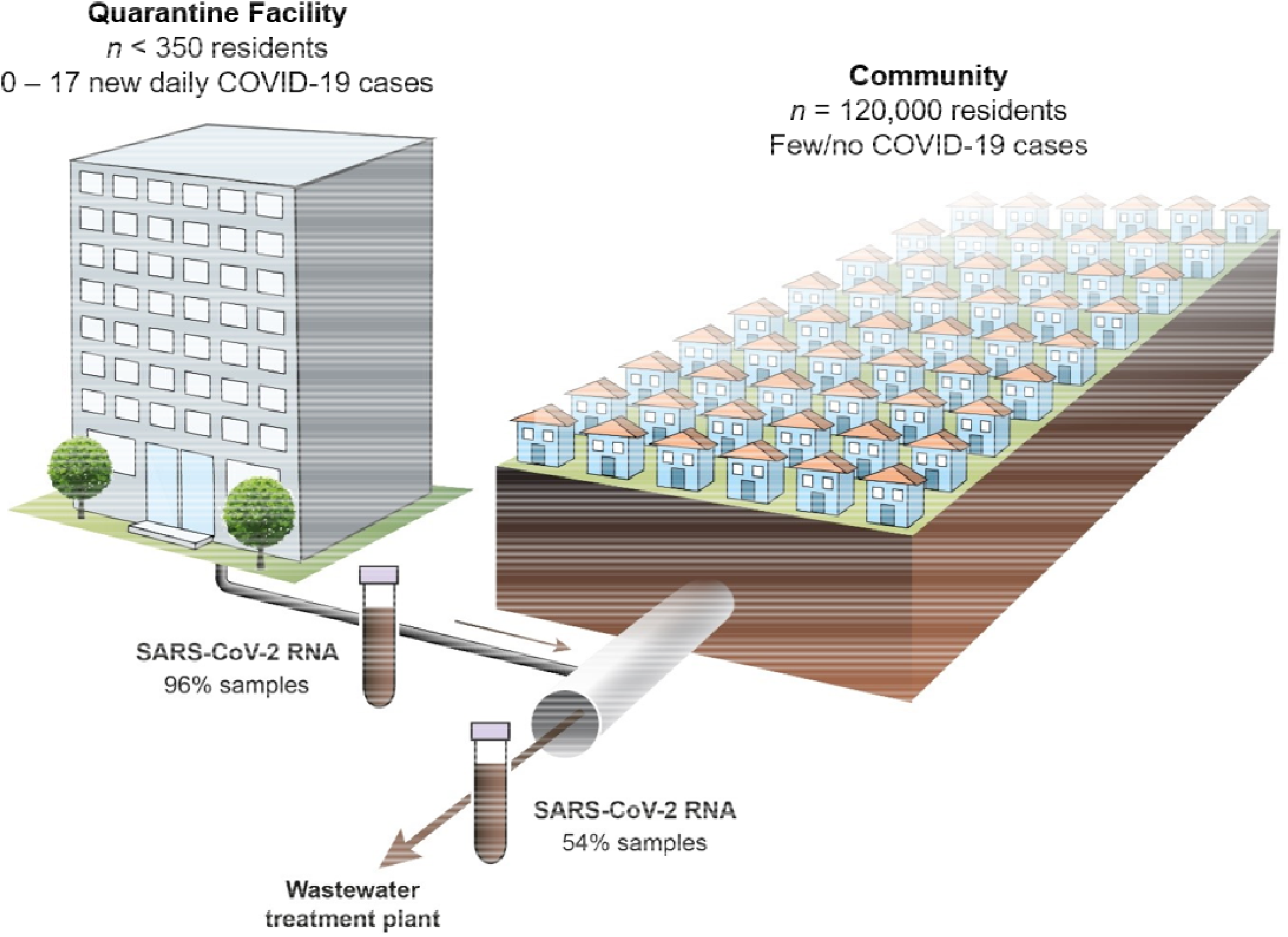

## 1. Introduction

Severe acute respiratory syndrome coronavirus 2 (SARS-CoV-2) has rapidly spread around the world since first detected in late 2019 (World Health Organisation 2020). For many countries, the virus has been extremely difficult to control and continues to circulate in the community (AJMC Staff 2021). SARS-CoV-2 RNA is shed into wastewater via faeces and respiratory excretions of symptomatic and asymptomatic individuals (Wölfel et al. 2020, Zhang et al. 2020). This means that wastewater-based epidemiology (WBE) can be used to monitor infections at the community scale (e.g. Lodder and Husman 2020, Peccia et al. 2020, Thompson et al. 2020). WBE complements other COVID-19 surveillance activities (i.e., diagnostic testing of mainly symptomatic individuals) and can operate at a much broader-scale (Bivins et al. 2020, Sims and Kasprzyk-Hordern 2020). WBE has been coupled with modelling to provide advanced warning (4-to 7-days) of an increase in cases (Peccia et al. 2020, Randazzo et al. 2020) and to estimate numbers of asymptomatic and/or undiagnosed cases (Chavarria-Miro et al. 2021, Wu et al. 2020b). WBE has been used at the commercial building or university campus scale to identify unknown cases, and better target public health interventions such as increased screening (Betancourt et al. 2021, Gibas et al. 2021, Harris-Lovett et al. 2021).

However, questions remain regarding the sensitivity of wastewater testing to detect infectious COVID-19 cases in communities where there no or limited cases (Cao et al. 2020, Foladori et al. 2020, Medema et al. 2020a, Pecson et al. 2021). It is currently unknown how many cases, per unit of population, are required for SARS-CoV-2 RNA to be reliably detected in municipal wastewater. In locations where the virus is widely circulating, detection in municipal wastewater may be difficult to estimate because undiagnosed or asymptomatic individuals may not be included in case counts. This scenario would result in the sensitivity of detection in wastewater being overestimated. Overestimation would potentially be exacerbated when the number of new cases per day, rather than total cases (i.e., incidence rather than prevalence) is used, given that individuals may continue to shed SARS-CoV-2 RNA for weeks following diagnosis (Cevik et al. 2021). While not ideal, daily incidence is the best proxy for total case load in many countries.

Aotearoa New Zealand adopted an COVID-19 elimination strategy with the intent of reducing the incidence of cases in the community to zero (e.g., Baker et al. 2020, Jefferies et al. 2020). Since 10 April 2020, all people entering New Zealand have been required to enter a government-run hotel-based managed isolation facility (MIF) upon arrival. All people are tested for COVID-19 on at least days 3 and 12, and if COVID-19 symptoms develop (New Zealand Government 2021). Most individuals who test positive are transferred to a single dedicated managed isolation and quarantine facility (MIQF) in Auckland. In many instances, their close contacts will be transferred at the same time. COVID-19 cases detected in the Auckland community, and their close contacts, are transferred from their home to the MIQF. This MIF system for border arrivals, along with a national lockdown and other measures, meant that by the beginning of June 2020, SARS-CoV-2 transmission was effectively eliminated in New Zealand (Baker et al. 2020). Since then, most New Zealand COVID-19 cases have been contained within one (MIQF) building. Interestingly, the MIFQ sits at the end of a wastewater pipe with no sewer upstream. The wastewater from the MIQF drains into a wider sewer pipe network with a resident population catchment of about 120,000. By testing paired daily influent wastewater samples from the sewer outside the MIQF and downstream sewer for the presence of SARS-CoV-2 RNA and comparing results to the known number of symptomatic and asymptomatic COVID-19 cases, we were able to calculate the sensitivity of WBE COVID-19 detection. Four shedding models were used to estimate the contribution of COVID-19 cases to the MIQF and downstream WWTP influent wastewater based on modelled infectious status.

## 2. Materials and Methods

### 2.1 Wastewater sampling

Time-weighted composite samples were collected from two sewer sites in the same wastewater catchment. At the MIQF, a temporary sewage maintenance access cover and ISCO autosampler (model 3710, Teledyne ISCO, Lincoln, NE) were installed in the sewer on 7 July 2020. The sampling point included intermittent pumped flow from two separate wastewater sumps on the premises. The sampler was programmed to collect 60 mL wastewater every 10 min per 24-hr period into a large plastic drum without refrigeration. The site was visited daily between 12:30 and 13:00 and, after stirring, a 1 L aliquot of the 24-hr wastewater composite was collected in a plastic bottle and transported on ice to the laboratory. At the inlet to the WWTP, relative to the associated interceptor of the MIQF, an ISCO autosampler (model 4700, Teledyne ISCO) was programmed to collect 200 mL wastewater every 30 min over 24 hr into a large plastic drum with refrigeration. The site was visited daily between 08:00 and 08:30 and, after stirring, a 1 L aliquot of the 24-hr composite was collected in a plastic bottle and transported on ice to the laboratory. On arrival at the laboratory, samples were processed within 18 hr of receipt (Sections 2.2 -2.5).

### 2.2 Quality controls

Feline infectious peritonitis virus (FIPV), an enveloped coronavirus, and murine norovirus (MNV), a non-enveloped virus, were used as external process controls to monitor virus recovery and to assess any RT-qPCR inhibition. Process control recoveries were not used to adjust the SARS-CoV-2 RNA concentration of the samples (as recommended by Kantor et al. 2021).

FIPV (ATCC® VR-990 WSU 79-1146) stocks were prepared, and 50% tissue culture infectious dose (TCID_50_)/mL determined, in Crandell-Rees Feline Kidney (CrFK) cells (ATCC® CCL-94). MNV stocks were prepared, and plaque forming units (PFU)/mL determined, in RAW 264.7 cells (Hewitt et al. 2009). Working stocks of 10^6^ TCID_50_/mL FIPV and 10^6^ PFU/mL MNV were prepared from the initial virus preparations by diluting in 2% Eagle’s Minimal Essential Media (Gibco, Waltham, MA). Aliquots were stored at −80 °C. Prior to virus concentration (Section 2.3), 2 µL of FIPV and MNV working stocks were added per 100 mL wastewater (e.g., 10 µL each of FIPV and MNV added to 500 mL). For each batch of samples processed, 10 µL each of FIPV and MNV working stocks were also added to 2.5 mL phosphate bu□ered saline (PBS), pH 7.2 and stored at 4 °C (for up to 18 hr). This was referred to as the FIPV/MNV in PBS process control.

A SARS-CoV-2 positive extraction quality control was prepared from a clinical specimen submitted for diagnostic testing (ESR, Porirua, NZ). The virus suspension was heat inactivated at 56 °C for 30 min, diluted 1/1000 in viral transport media, 200 µL aliquots prepared and stored at −80 °C until required for RNA extraction. The SARS-CoV-2 extraction control was quantified (as described below) and determined as approx. 30,000 genome copies (GC)/mL.

### 2.3 Viral concentration

Following seeding of FIPV and MNV into wastewater (Section 2.2), viruses from both liquid and solid fractions were recovered and concentrated using PEG precipitation. Wastewater (500 mL) was centrifuged at 10,000 x g for 20 min at 4 °C. The supernatant was transferred to a clean bottle and temporarily stored at 4 °C (maximum 2 hr). The solids were resuspended in a 1:8 (w/v) ratio of glycine buffer (0.05 M glycine, 3% beef extract), and pH adjusted to 9.0. The bottles were sonicated in an ultrasonic water bath for 2 min, and then placed on a horizontal shaker at 200 rpm for 30 min at room temperature. Subsequently, the glycine buffer mixture was centrifuged at 10,000 x g for 20 min at 4 °C and the supernatant combined with the initial, chilled, supernatant. The pH of the entire sample was adjusted to 7.0-7.2. Polyethylene glycol 6000 (Sigma-Aldrich, St. Louis, MO) and sodium chloride (Labserv, Auckland, New Zealand) were added to give final concentrations of 20% and 0.2 M, respectively, and gently mixed at room temperature until dissolved. The sample was then placed on a horizontal shaker and gently agitated at 70-80 rpm at 4 °C for a minimum of 2 h. The sample was centrifuged at 10,000 x g for 30 min at 4 °C and supernatant discarded. PBS, pH 7.2 was added to resuspend the pellet. The final ‘volume before processing’ to ‘volume after concentration’ ratio was approximately 200:1. The final volume was recorded and used in subsequent calculations to determine SARS-CoV-2 RNA GC/L wastewater, and to determine process control recoveries. Wastewater concentrates were stored at 4 °C (maximum 24 hr) until viral RNA was extracted.

### 2.4 RNA extraction

The High Pure Viral Nucleic Acid Extraction Kit (Roche Molecular Biochemicals Ltd., Mannheim, Germany) was used to extract viral nucleic acid from a 200 µL aliquot of each wastewater concentrate and the FIPV/MNV in PBS process control. One modification was made to the standard protocol; samples were centrifuged at 10,000 x g for 30 sec prior to addition to the filter. To monitor for RT-qPCR inhibition and/or poor RNA extraction, a 1:4 dilution of the concentrate and the FIPV/MNV in PBS process control was also prepared prior to RNA extraction. For each batch of RNA extractions, a positive (200 µL pre-aliquoted heat inactivated SARS-CoV-2 suspension) and negative (water) extraction control were used. Viral RNA was stored at 4 °C (maximum 1 hr) or at −80 °C until cDNA synthesis.

### 2.5 RT-qPCR

Two-step reverse transcription (RT)-qPCR assays were used for the detection of SARS-CoV-2, FIPV and MNV. Primers and probes are shown in Table S1. For all targets, viral cDNA (5 µL) was first generated from 2.5 µL RNA using Superscript^™^ III Reverse Transcriptase (Invitrogen) with specific reverse primers (Table S1) according to manufacturer’s instructions. RT reactions were carried out at 50 °C for 30 min followed by 95 °C for 4 min. The qPCR consisted of 5 µL cDNA, 10 µL PerfecTa ToughMix (Quantabio, Beverly, MA), appropriate primers and probe (Biosearch Technologies, Inc., Teddington, UK) and UltraPure DNase/RNase-free distilled water (DNase/RNase-free water, Invitrogen) to a final volume of 20 µL. Final primer/probe concentrations were 0.5 µM/0.25 µM (N-gene), 0.6 µM/0.15 µM (FIPV) and 0.4 µM/0.2 µM (MNV). The PCR cycling conditions for all three assays were 95 □ for 3 min, followed by 45 cycles of 95 □ for 15 secs and 60 □ for 30 secs. The N-gene qPCR assay was performed using the CFX96 Touch™ Real-Time PCR Detection System (Bio-Rad Laboratories, Hercules, CA) and the FIPV and MNV assays performed using CFX Connect™ (Bio-Rad). All assays were performed in hard shell, clear well, 96-well PCR plates (Bio-Rad), sealed with Microseal B (Bio-Rad). To increase the sensitivity of detection of SARS-CoV-2 RNA, each viral RNA extract from the wastewater concentrates was tested by RT-qPCR in quadruplicate for both extraction volumes (to give a total of eight reactions per sample). FIPV and MNV RT-qPCR assays were performed in duplicate, for both extraction volumes.

Controls for the N-gene RT-qPCR assay consisted of SARS-CoV-2 (positive) and DNase/RNase-free water (negative) extraction controls, non-template control (RT and qPCR) and reagent blank (qPCR). In addition, 5 µL (total 2 x10^2^ GC) N-gene synthetic DNA oligonucleotide standard (Dnature Diagnostics & Research Ltd, Gisborne, New Zealand) was included as a positive qPCR control (qPCR only). For FIPV and MNV, a FIPV/MNV in PBS process control (see Section 2.2), RNase/DNase-free water (negative extraction control), non-template water control (RT and qPCR), and reagent blank (qPCR) were used.

PCR cycle quantification threshold (Cq) values were determined in the Bio-Rad CFX Manager™ Software (Bio-Rad) after correcting for fluorescent drift and manually adjusting to a standardised threshold.

Synthetic SARS-CoV-2 RNA Fragment 1 Standard (National Institute of Standards and Technology, Gaithersburg, MD) was used to generate a RNA standard curve and to determine the limit of detection (LOD). RNA standard dilutions of 10^5^, 10^4^, 10^3^, 10^2^, 50, 25, 10, 5, 2 and 1 GC/reaction were prepared in DNase/RNase free water (Invitrogen) in DNA LoBind tubes (Eppendorf, Hamburg, Germany) to minimise RNA binding. The parameters of the standard curve derived RT-qPCR replicates of 10^5^ to 10 GC/reaction were slope, y-intercept, efficiency and R^2^ value of −3.580, 37.811, 90.3% and 0.991 respectively.

To determine the 95% LOD (GC/reaction), eight replicates of 10^3^, 10^2^, 50 and 25 GC/reaction, 15 replicates of 10 GC/reaction, and 20 replicates of 5, 2 and 1 GC/reaction were analysed. The RNA standard was detected in all replicates down to 10 GC/reaction (mean PCR Cq value 34.8 ± 0.4), and in 95% of the 5 GC/reaction replicates (mean PCR Cq value 36.1 ± 0.9). One and two GC/reaction were detected in approximately 50% RT-qPCR replicates (mean PCR Cq value 37.2 ± 0.7). The 95% LOD was 5 GC/reaction. This equated to 3.4 log_10_ SARS-CoV-2 GC/L (2500 GC/L) wastewater.

Results were expressed as positive (SARS-CoV-2 RNA detected) if SARS-CoV-2 RNA was detected in at least one RT-qPCR replicate (with a PCR Cq value ≤ 41), or negative (SARS-CoV-2 not detected). For the purposes of this study, SARS-CoV-2 RNA quantification was performed for wastewater samples when three or four RT-qPCR replicates were positive and where the mean PCR Cq value ≤ 36.1 (i.e., equivalent to 5 GC/reaction or greater), with a standard deviation (SD) of the PCR Cq values ≤1.0. For these samples, PCR Cq values were converted to GC/reaction using the standard curve, and then expressed as GC/L wastewater using the formula:

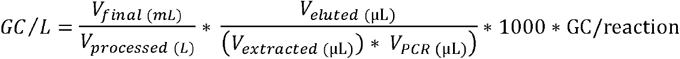

where V_final_ was the final volume of concentrate (mL), V_processed_ was the initial volume of wastewater processed (L), V_eluted_ was the volume following RNA extraction (µL), V_extracted_ was the volume of concentrate extracted (µL), and V_PCR_ was the volume of RNA used in the RT-qPCR reaction (µL). GC/L was then converted to log_10_ GC/L wastewater.

When three or four replicates were positive, but the calculated mean GC/reaction value was < 5 (i.e., < LOD per reaction), the SARS-CoV-2 RNA quantification (GC/reaction) was imputed as the LOD (i.e., 5) divided by the square root of 2 (Canales et al. 2018). This equated to 3.2 log_10_ GC/L wastewater for a concentration factor of 200. For samples that did not meet either of the above criteria, results were qualitatively presented as presence/absence detection differentiated by either 1 or 2 positive RT-qPCR replicates.

FIPV and MNV process control recoveries were calculated using the formula,

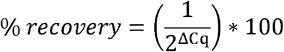

where ΔCq was the difference between the mean PCR Cq of the process control recovered in the sample and mean PCR Cq of the FIPV/MNV in PBS process control. Process control recoveries were considered acceptable for quality purposes if both were greater than 1%. By comparing the mean FIPV and MNV PCR Cq values of undiluted and 1:4 diluted samples, any RT-qPCR inhibition could be evaluated by observing a shift in PCR Cq values between the two samples. The 1:4 dilution would theoretically shift the Cq value by 2 (i.e., a diluted sample gives a higher PCR Cq value when there is no inhibition).

### 2.6 Epidemiological data

Epidemiological data on notified COVID-19 cases (symptomatic or asymptomatic) were obtained from the EpiSurv Surveillance database (https://surv.esr.cri.nz/episurv). These included dates for arrival in the country, transfer to and from the MIQF, onset of illness (if symptomatic), and collection of clinical samples for diagnostic purposes. Based on these data, we used four models to estimate the shedding of SARS-CoV-2 RNA into wastewater based on modelled infectious status:

- **Model 1: Total cases** - A count of all COVID-19 cases irrespective of their likely infectious status (conservative overestimate).
- **Model 2: Infectious cases** - A count of likely infectious COVID-19 cases with a constant shedding pattern beginning three days before reported (symptomatic cases) or imputed (asymptomatic cases, see below) disease onset through to nine days after onset.
- **Model 3: Relative infectious cases** - A count of likely infectious COVID-19 cases with a proportional shedding pattern based on He et al. (2020) (Fig. 1), with peak shedding on the day of reported (symptomatic cases) or imputed (asymptomatic cases, see below) disease onset, and proportionately less before and afterwards. Due to the calculation, the relative infectious case number was not restricted to whole numbers.
- **Model 4: New daily cases -** A count of new cases per day (i.e., daily cases), based on reported (symptomatic cases) or imputed (asymptomatic cases, see below) onset of disease.

**Fig. 1.**
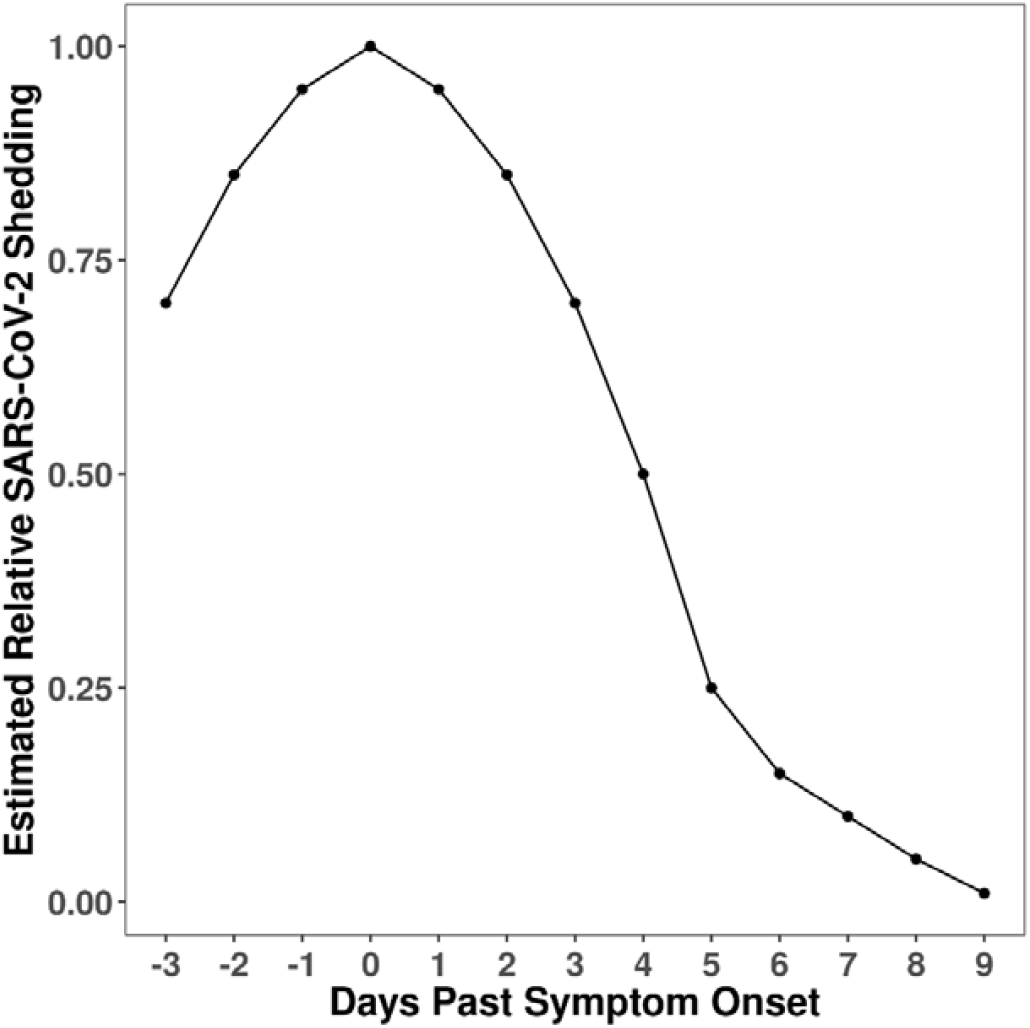
Model to estimate viral shedding relative to estimated symptom onset date (model 3). Peak shedding at symptom onset date (or estimated onset date), with proportionally less for 3 days prior, and 9 days after, symptom onset date. Model 3 was adapted from He et al. (2020).

Estimating the degree of infectiousness for individual cases likely provides more realistic estimates of the number of infected people shedding SARS-CoV-2 RNA to wastewater than the total cases in the catchment (Walsh et al. 2020). We assumed that peak shedding for asymptomatic cases occurred on the date that a positive nasopharyngeal swab sample was collected. We therefore used this sampling date as a proxy “onset date” and that the shedding profile was the same as symptomatic cases.

Often, SARS-CoV-2 RNA contributions made by shedding cases would not have been detected until the following day, due to the residence time of wastewater and the nature of the 24-hr sampling scheme. For example, SARS-CoV-2 RNA shed from a MIQF resident at 2pm on Monday would appear in Tuesday’s 24-hr composite samples collected at the MIQF and WWTP. The date of wastewater samples therefore reflects the contributions from the previous 24 hours (i.e., date is the day the 24-hr composite sampling ended and collected). Dates relating to occupation and shedding density contribution have been adjusted to reflect the date that wastewater was collected.

### 2.7 Statistical analysis

Mean (± bootstrap 95% confidence intervals (CI)) process control recovery efficiencies were calculated. Logistic regression was used to model the relationship between the number of COVID-19 cases/case-equivalents putatively shedding at the MIQF, as cases per 100,000 population, and detection of SARS-CoV-2 RNA in the paired MIQF and WWTP wastewater samples. The contributions of asymptomatic and symptomatic COVID-19 cases/case-equivalents putatively shedding at the MIQF (per 100,000 population) were regarded in additive models to examine the relative contribution of asymptomatic cases to WWTP wastewater samples. All statistical analyses were performed in R Statistical Computing Software version 4.0.3 (R Core Team 2020) with Tidyverse (Thompson et al. 2020) used to produce graphs.

## 3. Results

### 3.1 Cases in managed isolation and quarantine (MIQF)

From 9 July to 6 November 2020, there were a total of 335 COVID-19 cases at the MIQF. Of the 335 total cases, 212 were symptomatic and 123 were asymptomatic. Symptomatic cases had an illness onset which ranged from 30 days before arriving at the MIQF to 3 days after arrival, with an average onset of illness 3.3 days before arrival at the MIQF. Overall, COVID-19 cases stayed at the MIQF from 5 – 52 (mean of 13) days (Fig. 2). During the four-month study, one cluster of 179 community cases, mostly residing in the WWTP catchment being studied, was reported. All cases were rapidly identified (onset of illness 31 July – 11 September 2020), with 155 cases being transferred to the MIQF between 11 August and 11 September (Fig 2). On 29 August 2020, the total number of cases at the MIQF peaked at 107. This was equivalent to 40 infectious cases (model 2) or 16 relative infectious cases (model 3), and 17 new daily cases (model 4). There were no other known COVID-19 cases outside of the MIQF during this study period. At any one time, there were 5 – 107 total cases (model 1) (equivalent to 4.1 – 89.2 cases per 100,000 population), 2.5 – 50.3 infectious cases (model 2) or 0.2 – 15.1 relative infectious cases (model 3). New daily cases at MIQF (model 4) ranged from a total of 0 – 17 (or 0 – 14.1 per 100,000 population), with an average of two new cases per day. There were between 41 and 121 (average 93) staff, security, or other non-guests onsite at the MIQF each day (none of them cases), giving a total population at the MIQF each day of about 300-500 individuals who potentially contributed to the wastewater.

**Fig. 2.**
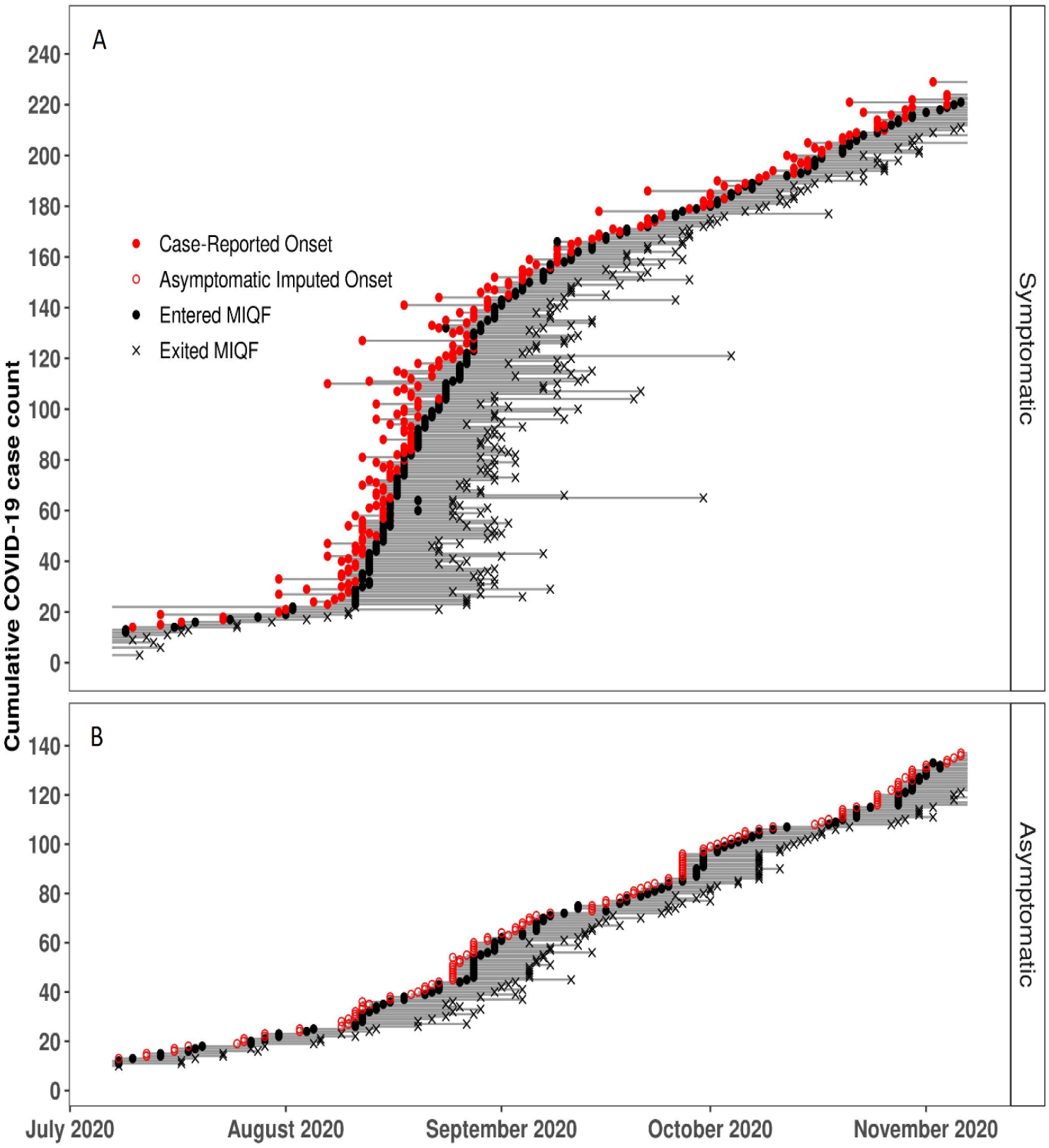
History of each symptomatic (Panel A) and asymptomatic (Panel B) COVID-19 case at the managed isolation and quarantine facility (MIQF). Closed red circles indicate the date reported as symptom onset, open red circles indicate the imputed symptom onset date for asymptomatic cases. Closed black circles indicate the date the case entered the MIQF. Grey lines indicate dates each case was present in MIQF. Black crosses indicate the date the case exited the MIQF.

### 3.2 Process control recovery from wastewater

Two samples from the MIQF, collected on 19 August and 25 September 2020, had unusual low pH/visual properties with low FIPV recovery (0.3% and 0.5% respectively). This may have been a result of excess acidic commercial cleaning products being used at the MIQF. Data from the four samples (two MIFQ and two WWTP wastewater) collected on these two dates were excluded from further analyses. Data from the remaining 113 paired samples (*n* = 226) were used for subsequent analyses.

Process control recoveries (%) (mean ± 95% CI) from MIQF wastewater samples were 25.4% (22.5, 28.5) for FIPV and 20.8% (18.1, 23.8) for MNV (Fig. S1a). Recoveries from WWTP wastewater samples were 20.7% (16.7, 25.1) for FIPV and 33.9 % (30.2, 38.2) for MNV (Fig. S1a). In samples diluted 1:4 prior to RNA extraction, FIPV and MNV recovery data showed that, overall, there was no significant RT-qPCR inhibition detected in the MIQF or WWTP wastewater samples (Fig. S1b).

### 3.3 Detection and quantification of SARS-CoV-2 RNA in wastewater

SARS-CoV-2 RNA was detected in 95.6% (108/113) of the MIQF wastewater samples. Mean (± SD) PCR Cq values for samples ranged from 26.2 (± 0.1) to 37.9 (± 0.4) (Fig. 3a). For the majority (84.3%, 91/108) of samples where SARS-CoV-2 RNA was detected, all four RT-qPCR replicates were positive. SARS-CoV-2 RNA concentrations in MIQF wastewater ranged from 3.2 to 6.0 log_10_ GC/L. Non-detection of SARS-CoV-2 RNA in MIQF wastewater occurred on 1 July, 25-27 July, and 3 August 2020.

**Fig. 3.**
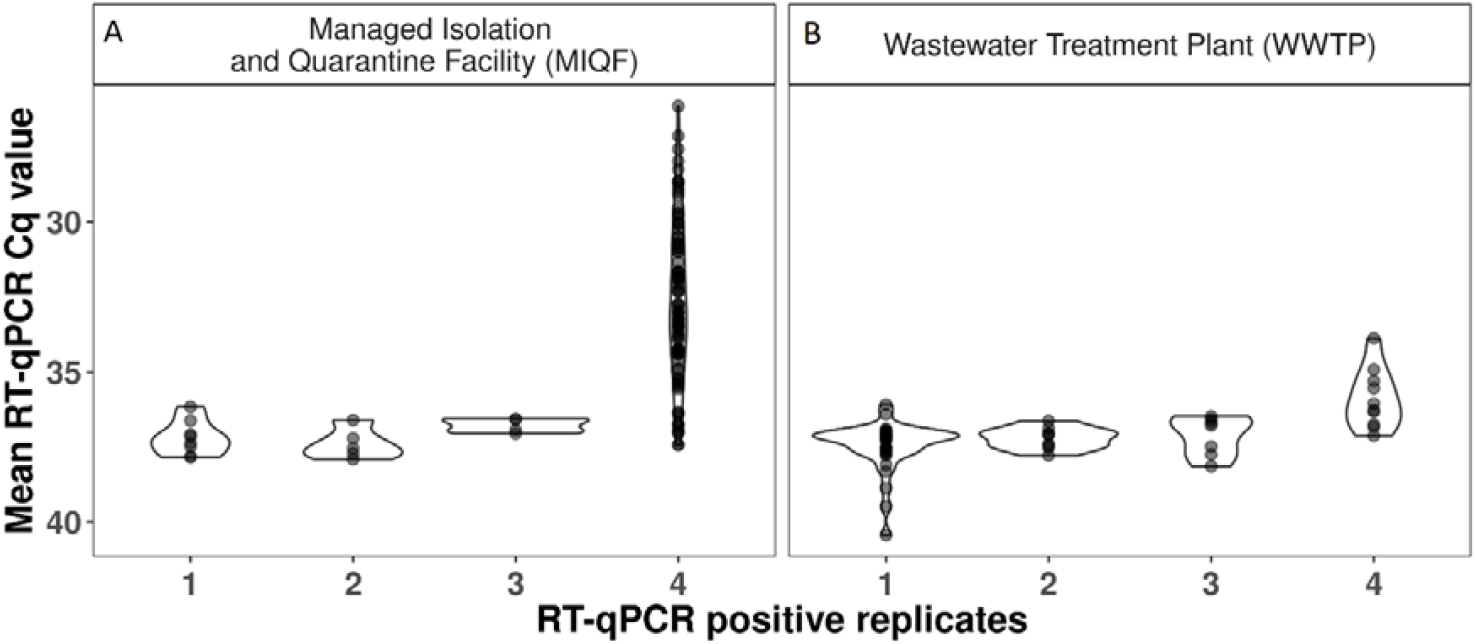
Number of RT-qPCR replicates positive for SARS-CoV-2 RNA in the MIQF (Panel A) and WWTP wastewater (Panel B) samples. Mean RT-qPCR Cq values are shown with an inverted y-axis (as lower Cq values represent higher viral concentrations).

SARS-CoV-2 RNA was detected in 54.0% (61/113) of the WWTP samples. Of the 61 samples where SARS-CoV-2 RNA was detected, 16.4% (10/61) had four positive RT-qPCR replicates. For these ten samples, mean (± SD) PCR Cq values ranged from 33.9 (± 0.7) to 38.1 (± 1.2) (Fig. 3b), with SARS-CoV-2 RNA concentrations that ranged from 3.3 to 3.8 log_10_ GC/L wastewater from the WWTP influent. For SARS-CoV-2 RNA detections with 1 to 3 positive RT-qPCR replicates, the mean (± SD) PCR Cq value was 37.3 ± 0.8 (range 35.6 to 40.4). Notably, over half of the positives (55.7%, 34/61) had only 1 of 4 positive RT-qPCR replicates (Fig. 3b).

SARS-CoV-2 RNA was not detected in daily samples from three other Auckland municipal WWTPs, serving a total of ca. 1.2 million population, expect for a few detections (data not shown) in the period of the community cluster in August and early September. This reflects the fact that the area (Auckland) was COVID-19 free during the study and provides confidence that the number of COVID-19 cases was very well quantified by measuring those people in the MIQF. Positive detections in the wastewater therefore were solely due to SARS-CoV-2 RNA input from the MIQF and so the data can be used to quantify detection sensitivity.

### 3.4 Relationship between COVID-19 cases and wastewater analysis

As may be expected, quantifiable SARS-CoV-2 RNA concentrations at the WWTP occurred most often when there were the highest number of individuals potentially shedding SARS-CoV-2 RNA (Fig. 4a and 4b). In addition to simply counting COVID-19 total cases, shedding was modelled on the more restricted shedding assumptions described above.

**Fig. 4.**
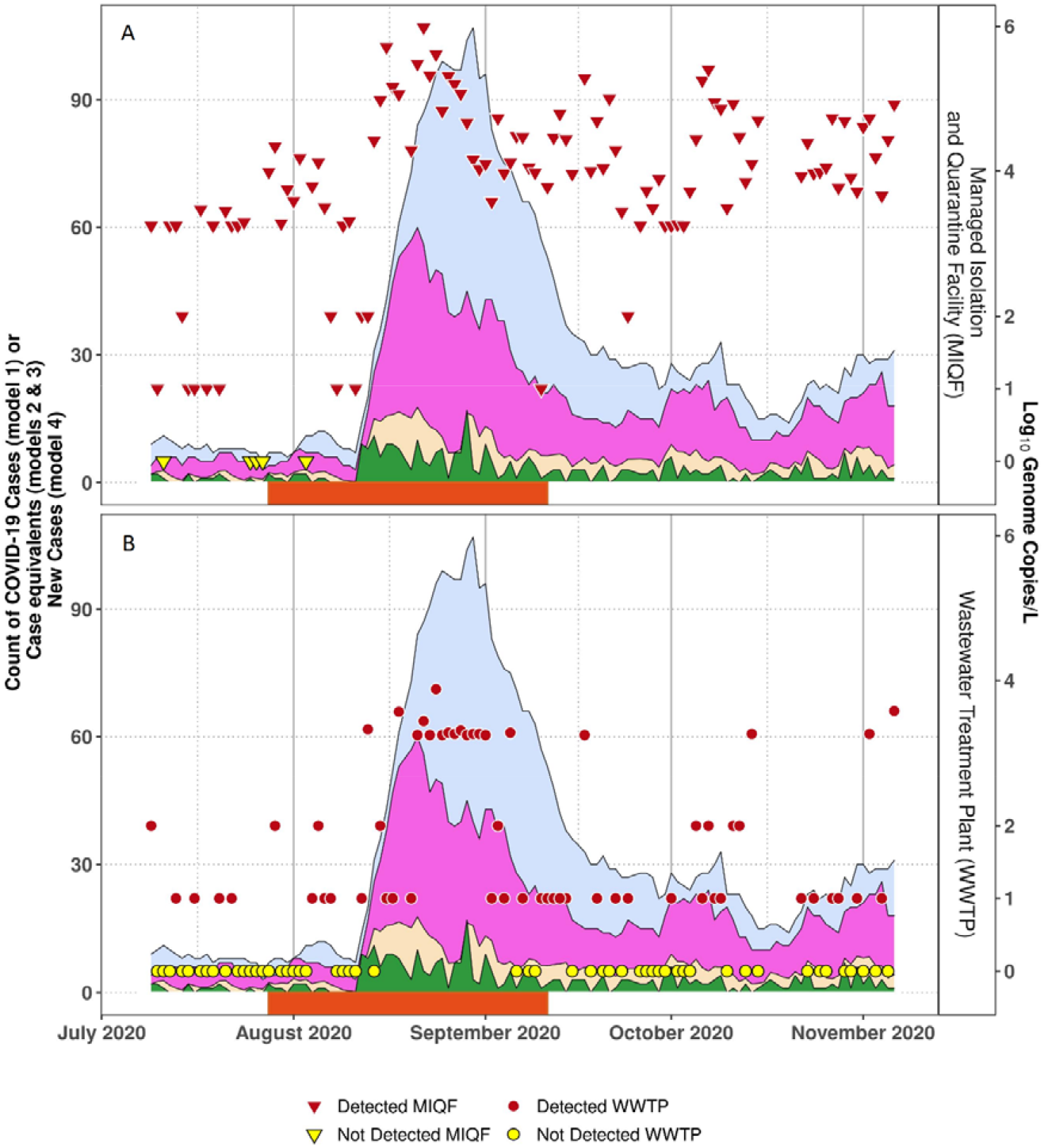
Detection of SARS-CoV-2 RNA in wastewater collected at the MIQF (Panel A) and at WWTP (Panel B). Red triangles indicate MIQF positive samples, yellow triangles indicate MIQF negative samples, red circles indicate WWTP positive samples, and yellow circles indicate WWTP negative samples. On the y-axis, log_10_ genome copies/L of 1 or 2 have been imputed to represent the 1 or 2 positive RT-qPCR replicates. Blue shading indicates COVID-19 cases at the MIQF on given day (model 1), purple shading estimated infectious cases (model 2), beige shading estimated relative infectious cases (model 3), and green shading new daily cases (model 4). The period when community cases were identified (28 July to 11 September 2020) is indicated by orange bar on x-axis.

When SARS-CoV-2 RNA was detected in the MIQF wastewater (*n* = 108), there were 6 - 107 total cases (i.e., 5 – 89 per 100,000 population) (model 1). This is equivalent to 3 −; 60 (i.e., 2.5 – 50 per 100,000 population) infectious cases (model 2) and 0.2 – 9.1(i.e., 0.2 – 7.6 per 100,000 population) relative infectious cases (model 3). Non-detections of SARS-CoV-2 RNA in the MIQF wastewater were associated with 5 – 11 total MIQF cases (i.e., 4 – 9 per 100,000 population) compared to at least 6 total cases when SARS-CoV-2 RNA was detected. The MIFQ wastewater samples with SARS-CoV-2 RNA concentrations more than 4.0 log_10_ GC/L were collected on days when there were more cases (i.e., 7 – 107 cases (i.e., 5.8 – 89.2 per 100,000 population), compared to 0 - 60 cases (i.e., 0 – 50 per 100,000 population) depending on the shedding assumptions (Table 1).

**Table 1.** Estimated COVID-19 case equivalents at the MIQF when SARS-CoV-2 RNA was either not detected, detected at ≤4.0 log_10_ GC/L or detected at >4.0 log_10_ GC/L wastewater collected at the MIQF (*n* = 113).

| SARS-CoV-2 RNA<br>in MIQF wastewater | No of<br>samples | Average number of COVID-19 cases<br>at the MIQF (Minimum - Maximum) |  |  |  |
| --- | --- | --- | --- | --- | --- |
|  |  | Total cases<br>Model 1 | Infectious cases<br>Model 2 | Relative<br>infectious cases<br>Model 3 | New daily cases<br>Model 4 |
| Not detected | 5 | 8.2 (5 - 11) | 5.2 (3 - 7) | 1.7 (0.8 – 3.0) | 0.8 (0 - 2) |
| Detected ($\leq 4.0 \log_{10}$ GC/L) | 56 | 21.1 (6 - 83) | 12.8 (3 - 43) | 4.2 (0.2 – 12.2) | 1.8 (0 - 9) |
| Detected ($> 4.0 \log_{10}$ GC/L) | 52 | 49.5 (7 - 107) | 27.4 (4 - 60) | 8.0 (2.0 – 17.7) | 3.9 (0 - 17) |
| Total | 113 | 33.6 (5 - 107) | 19.2 (3 - 60) | 5.8 (0.2 – 17.7) | 2.7 (0 - 17) |

It is clear from the data that detecting SARS-CoV-2 RNA at the WWTP was more likely when there were at least 8 total cases (i.e., using model 1) (Table 2). This threshold reduced to 4 cases using model 2 and one cases using model 3. SARS-CoV-2 RNA detections at the WWTP, particularly those with three or four RT-qPCR replicates, were associated with an increase in the number of total (model 1), infectious (model 2), relative infectious (model 3), or new daily (model 4) cases in the catchment (Table 2).

**Table 2.** Estimated COVID-19 case equivalents at the MIQF when SARS-CoV-2 RNA was either not detected, detected in 1 or 2 RT-qPCR replicates, or detected in 3 or 4 RT-qPCR replicates at the WWTP (*n* = 113).

| SARS-CoV-2 RNA<br>in WWTP wastewater | No of<br>samples | Average number of COVID-19 cases<br>at the MIQF (Minimum - Maximum) |  |  |  |
| --- | --- | --- | --- | --- | --- |
|  |  | Total cases<br>Model 1 | Infectious cases<br>Model 2 | Relative<br>infectious cases<br>Model 3 | New daily cases<br>Model 4 |
| Not detected | 52 | 20.1 (5 – 71) | 12.6 (3 - 28) | 4.0 (0.2 – 15.0) | 1.8 (0 - 11) |
| Detected: 1 or 2 replicates | 42 | 32.1 (8 - 83) | 19.6 (4 – 57) | 6.0 (1.0 – 15.9) | 2.5 (0 - 9) |
| Detected: 3 or 4 replicates | 19 | 75.0 (18 - 107) | 37.2 (10 - 60) | 10.3 (2.0 – 17.7) | 5.7 (0 - 17) |

Logistic regression models were fitted to data excluding and including the period of the community COVID-19 cluster for each shedding model (Fig. 5). When the data during the community cluster were excluded, increases in relative infectious cases and new daily cases, was not significantly associated with SARS-CoV-2 RNA detection at the WWTP (df = 67, model 3: p = 0.113; model 4: p = 0.372). Case load and infectious cases were significantly associated with an increase in SARS-CoV-2 RNA detection at the WWTP (df = 67, model 1: p = 0.012, model 2: p = 0.033) (Table S2).

**Fig. 5.**
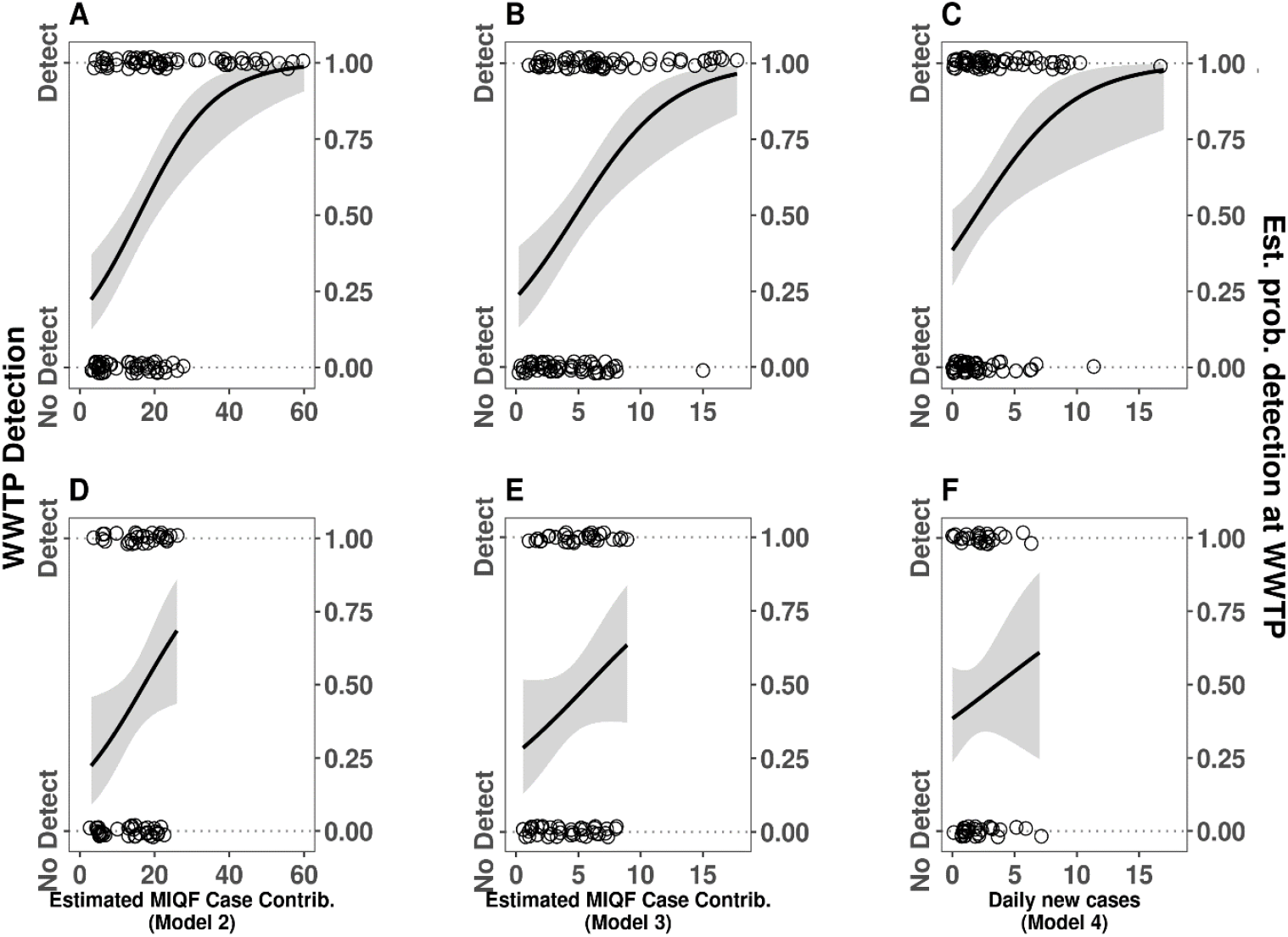
Logistic regression of the probability of detecting SARS-CoV-2 RNA at the WWTP, estimated using A) MIQF infectious cases (model 2) for the whole study period; B) MIQF relative infectious cases (model 3) for the whole study period; C) New daily cases (model 4) for the whole study period; D) MIQF infectious cases (model 2) excluding the community cluster data; E) MIQF relative infectious cases (model 3) excluding the community cluster data; F) New daily cases (model 4) excluding the community cluster data.

However, when the community cluster data was included, increasing cases for all models were associated with an increased probability of detection at the WWTP (df = 112, models 1, 2 & 3: p<0.001, model 4: p = 0.003) (Table S2).

The proportion of symptomatic to asymptomatic cases at the MIQF varied with SARS-CoV-2 RNA detections at the WWTP (Fig. 6). The interquartile range (IQR) of ratios was higher in the 19 WWTP samples with three or four positive RT-qPCR replicates (IQR = 2.5 – 5.8 symptomatic cases to every asymptomatic case) than for the 42 WWTP samples with only one or two positive RT-qPCR replicates (IQR = 0.78 – 2.4 symptomatic cases to every asymptomatic case) and for the 52 WWTP samples where SARS-CoV-2 RNA was not detected (IQR = 0.71 – 1.8 symptomatic cases to every asymptomatic case).

**Fig. 6.**
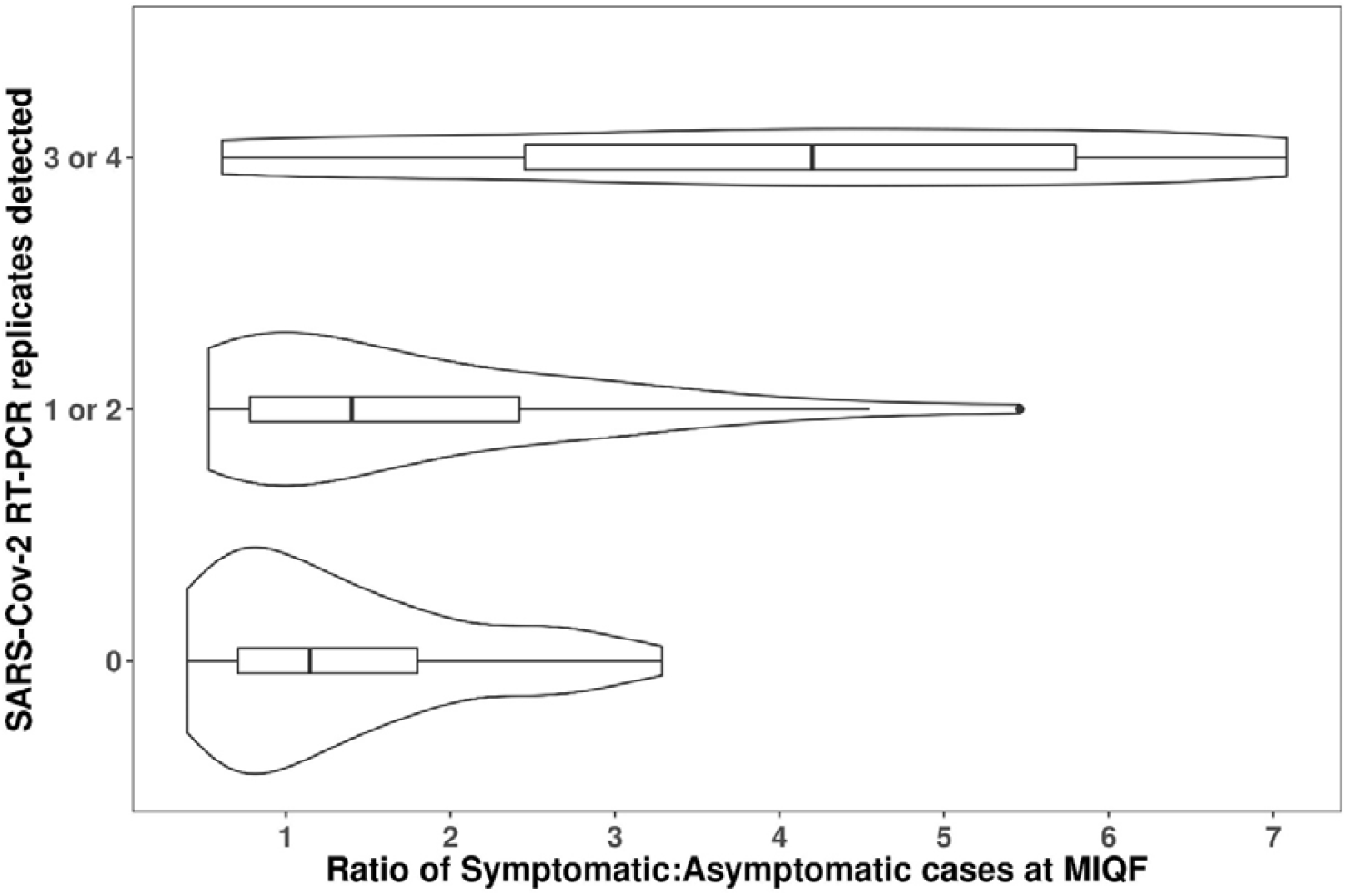
The proportion of symptomatic cases to asymptomatic cases (model 1) at the MIQF by the number of RT-qPCR replicates positive for SARS-CoV-2 RNA at the WWTP.

Logistic regression models with terms for symptomatic and asymptomatic case contributions for infectious cases (model 2) and relative infectious cases (model 3), and new daily cases (model 4) were significantly associated with an increase in probability of detection of SARS-CoV-2 RNA at the WWTP for symptomatic cases (df = 112, model 2 & 3: p = 0.002, model 4: p = 0.008) (Supp. Table S2). However, asymptomatic cases were not significantly associated with an increase in probability of detection of SARS-CoV-2 RNA at the WWTP for any shedding model (df = 112, model 2: p = 0.488, model 3: p = 0.661, model 4: p = 0.461) (Supp. Table S2). This indicates that asymptomatic cases are likely to be shedding less detectable SARS-CoV-2 RNA into wastewater.

## 4. Discussion

In this study, by comparing actual case counts with models estimating number of infectious individuals, we have shown that SARS-CoV-2 RNA can be detected in wastewater when relatively few infectious individuals are contributing to the sample. For example, detection probability was 87% when the infectious case prevalence (model 3) was 0.01% (i.e,.10 infectious cases per 100,000 population). While SARS-CoV-2 RNA can be detected when there is only one infectious case in 100,000 population, the predicted probability was lower at 29%. Hence, in communities with very low numbers of COVID-19 cases, false negative results may occur. This false negative result may occur at the sample scale, because the amount of viral RNA within the sample is below the limit of detection. Alternatively, the sample could be a true negative (i.e., no SARS-CoV-2 RNA is present in the sample) but be a false negative at the community scale because the sample collected did not capture the intermittent and low levels of SARS-CoV-2 RNA in the wastewater system. Even if sampling could somehow test all the wastewater in a community, shedding from infectious individuals whose residence is served by a septic tank or who are otherwise not connected to the wastewater system will also not be detected. These factors must be considered when interpreting and communicating results to public health authorities.

### WBE sensitivity estimates

Previous estimates of the sensitivity of SARS-CoV-2 RNA detection in wastewater have varied widely and few have considered the effect that variable shedding patterns over the course of an infection will have on estimates. Hart et al. (2020) suggest that, depending on local conditions, a single COVID-19 case would be theoretically detectable using WBE from a catchment of 100 (1% infection rate, least sensitive) to 2,000,000 (0.00005% infection rate, most sensitive) people, based on excretion rates, wastewater flow rates and detection limit of 10 GC/mL (i.e., 10^4^ GC/L) wastewater. Our data tested these estimates and found that detection with 0.00005% prevalence would be highly unlikely, whereas detection with a 1% prevalence would be highly likely. Indeed, detection with prevalence of 0.005% was possible, but was more reliable (90-100%) at a 0.03% or higher prevalence (i.e., 30 total cases in 100,000 population). The sensitivity reported in our study is similar to that reported by Hata et al. (2021) who concluded that, while there was a possibility that SARS-CoV-2 RNA could be detected in wastewater when the number of cases was less than 1 per 100,000 (0.001%), detections were more likely when COVID-19 cases exceeded 10 per 100,000 population (0.01%). Medema et al. (2020b) reported somewhat higher sensitivity, with positive detections in wastewater occurring when the prevalence of COVID-19 cases was higher than 5 cases per 100,000 population (0.005%) (Table 3). Other studies (e.g., Baldovin et al. 2021, Hong et al. 2021) have reported lower sensitivity. Differences in sensitivity between studies may be caused by variation in number of asymptomatic and pre-symptomatic individuals, and COVID-19 testing rates, in different localities. As the number of undetected COVID-19 cases contributing to wastewater increases, precision of sensitivity estimates reduces. Most previous studies were based on reported cases, which are assumed to underrepresent actual infections in communities with widespread community transmission.

**Table 3:**
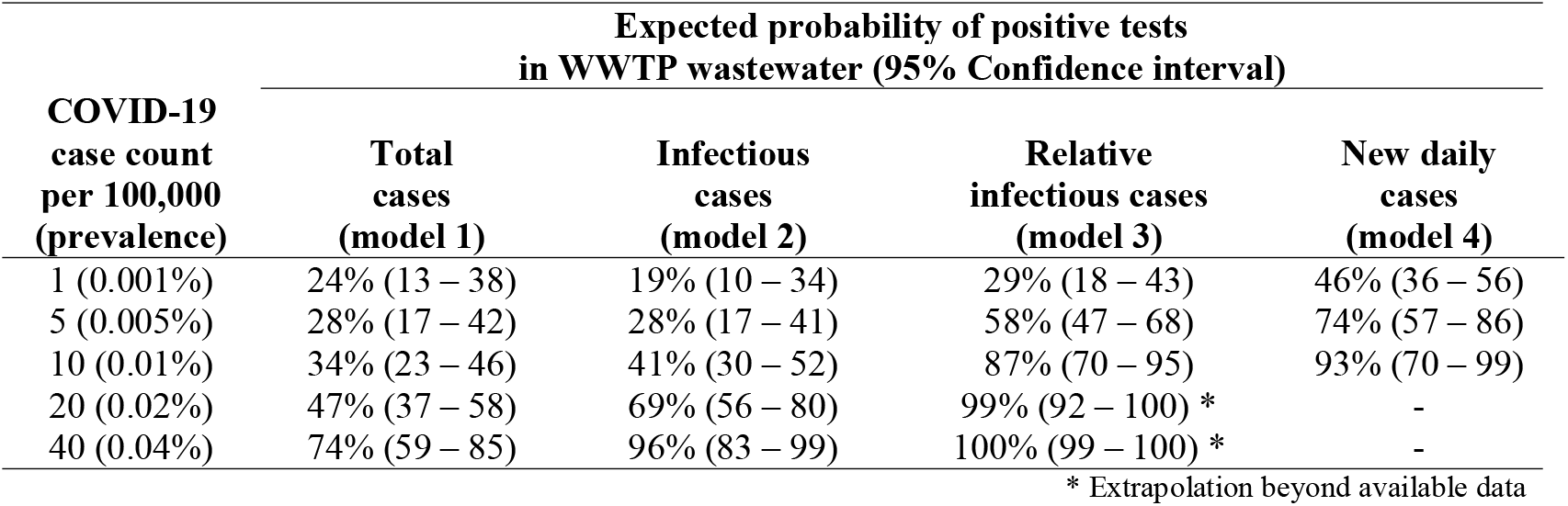
Predicted probabilities of SARS-CoV-2 RNA detection at the WWTP by selected equivalent case counts (per 100,000 population) from logistic regression models fitted with community cluster data.

### Use of shedding models

The amount of shedding by individuals is known to vary person to person, and over time (Hoffmann and Alsing 2021). In this study, some individuals stayed at the MIQF for over 30 days (average 13 days), during which time the amount of SARS-CoV-2 RNA shed via faeces and respiratory excretions likely varied from high to undetectable levels Using total COVID-19 cases as an indicator of SARS-CoV-2 RNA shedding per person provides a conservative estimate of the sensitivity of detecting SARS-CoV-2 RNA in wastewater. This is because individuals towards the end of their infectious period, and potentially shedding at low levels, are counted equally with people at the start of their infectious period. Both models 2 and 3 assumed that viral shedding was minimal nine days after illness onset (Cevik et al. 2021). While SARS-CoV-2 RNA continues to be shed for prolonged periods in some individuals (Gupta et al. 2020), which could have resulted in a sensitivity overestimate in our study, the impact of this is likely to be minimal, as almost all cases left the MIQF (and most presumably from the larger WWTP catchment) within 12 days of reported illness onset. Models 3 and 4 provided similar estimates of detection probability. This similarity likely reflects that detection (and movement into MIQF) for most cases occurred in the early stages of their infection. Unsurprisingly, the probability of SARS-CoV-2 RNA detection at the WWTP increased as case numbers rose. This was evident during the period of community cases within the study catchment, when SARS-CoV-2 RNA was detected with quantifiable concentrations at the WWTP. Over the course of this study, the number of new daily cases (model 4) ranged from 0 to 17, with an average of two new cases per day. Using new daily cases to estimate a positive detection of SARS-CoV-2 RNA therefore had little support beyond 10 new daily cases due to the low case prevalence during the study period (Fig. 5c).

### Shedding by asymptomatic cases

Our data suggest that symptomatic individuals may shed a higher amount of SARS-CoV-2 RNA into wastewater than asymptomatic individuals (Fig. 6). Wastewater testing is potentially one of the best ways to detect asymptomatic cases in a community, as they are otherwise unlikely to be tested unless they are close contacts of symptomatic cases. During our study, asymptomatic case load was not significantly associated with increased SARS-CoV-2 RNA detection in wastewater for any shedding models. It is possible that asymptomatic cases in our study may have been tested longer after infection than symptomatic cases, many of whom were tested immediately upon developing symptoms. Studies specifically investigating differences in shedding between symptomatic and asymptomatic individuals, particularly in faeces, would be valuable. We are aware of only a few studies where SARS-CoV-2 RNA has been detected in the faeces of asymptomatic individuals (e.g., Li et al. 2020, Yuan et al. 2021).

### Near-source tracking

As part of this study, we have showed that at a building level, wastewater analysis has a very high probability of detecting SARS-CoV-2 RNA shed from COVID-19 cases, even when total infectious COVID-19 cases are few. This wastewater ‘near-source tracking’ has been used to target surveillance where it is most needed (Medema et al. 2020a) such as in university campuses and schools (Crowe et al. 2021), elderly care facilities and hospitals (Davó et al. 2021, Spurbeck et al. 2021), and households (Döhla et al. 2020). Similar to the concept of the MIQF used in our study, Hong et al. (2021) used a semi-controlled community, namely a hospital with COVID-19 cases in Jeddah, Saudi Arabia, to examine the detection sensitivity of WBE. They calculated sensitivity of ⍰253 positive cases per 10,000 population (2.5% prevalence) for reliable detection in wastewater collected via a grab-sample. Baldovin et al. (2021) sampled localised urban wastewater lines in Padua, Italy during a time of high COVID-19 case numbers. The authors calculated a detection sensitivity of 1 case per 531 hospitalised inhabitants (0.19% prevalence) for reliable detection.

### Sampling frequency

Daily sampling, as undertaken in this study, provides the best understanding of potential viral RNA shedding in the community and the earliest response opportunity. Based on our study, detecting 0.8 to 4.2 cases in 100,000 is possible, although shedding dynamics, and wastewater factors such as dilution and distance of cases from the sampling site, means that the likelihood of detection in a single wastewater sample might only be 20-50%. As daily testing may not be practical, investigations around the optimal frequency of wastewater sampling in a low prevalence setting would be a fruitful area of research.

## Conclusions

By quantifying the detection limit of SARS-CoV-2 RNA in wastewater where the number of contributing symptomatic and asymptomatic COVID-19 cases was known, and through modelling viral RNA shedding, this study demonstrates that even with very low numbers of infected individuals in a WWTP catchment there is a high probability (>90 % depending on shedding assumption model) of detecting SARS-CoV-2 RNA in wastewater. As such, wastewater can be used as both an early warning system of community outbreaks and for longer term surveillance. However, it is important to carefully consider how cases are quantified/modelled, and the relative contributions of symptomatic and asymptomatic individuals when estimating sensitivity and interpreting results of WBE studies of SARS-CoV-2.

## Data Availability

All data is included in the paper

## Acknowledgements

The authors would like to thank residents and staff at the MIQF and WWTP catchment for their wastewater inputs, and staff who collected samples and helped with logistics. Carmen Vettori, Donna Marie Warren and Mary Brunsdon (ESR) assisted with sample accessioning and laboratory support services, Alex Eustace, Amy Bradshaw and Tammy Waters (ESR) helped with sample processing and/or analysis, Beverly Horn and Jenny Ralston (ESR) provided assistance with statistical analysis and graphics respectively. Magda Dunowska (Massey University, New Zealand) provided the feline infectious peritonitis virus (FIPV), ATCC VR-990 strain WSU 79-1146 and designed the primers. Pradip Gyawali (ESR) designed the FIPV probe. Murine norovirus was kindly provided by Prof Herbert Virgin (Washington University School of Medicine, MO, US). Watershed Engineering Ltd conducted mapping of the sewershed and developed estimates of resident population. Liza Lopez (ESR) extracted case data from EpiSurv - a secure web-based application based on the Surveillance Information New Zealand (SurvINZ) architecture. The authors thank the team that maintain EpiSurv and those who have uploaded case data. We also appreciate comments on an earlier version from Erasmus Smit, Gerard Sonder and Phil Carter (ESR).

---

**Supplementary Table S1.**
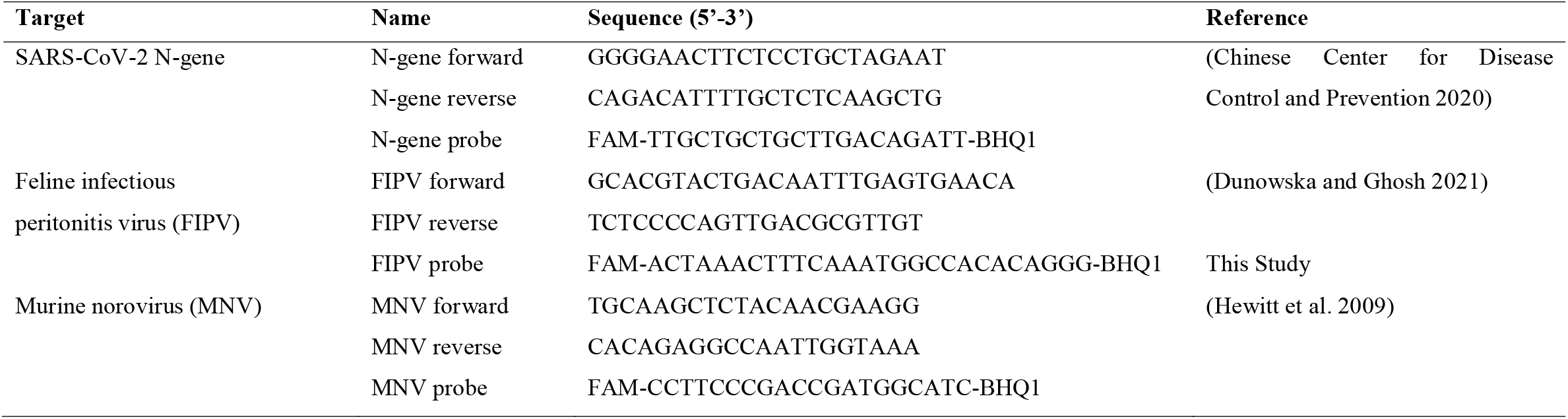
Primers and probes used for this study.

**Supplementary Table S2.**
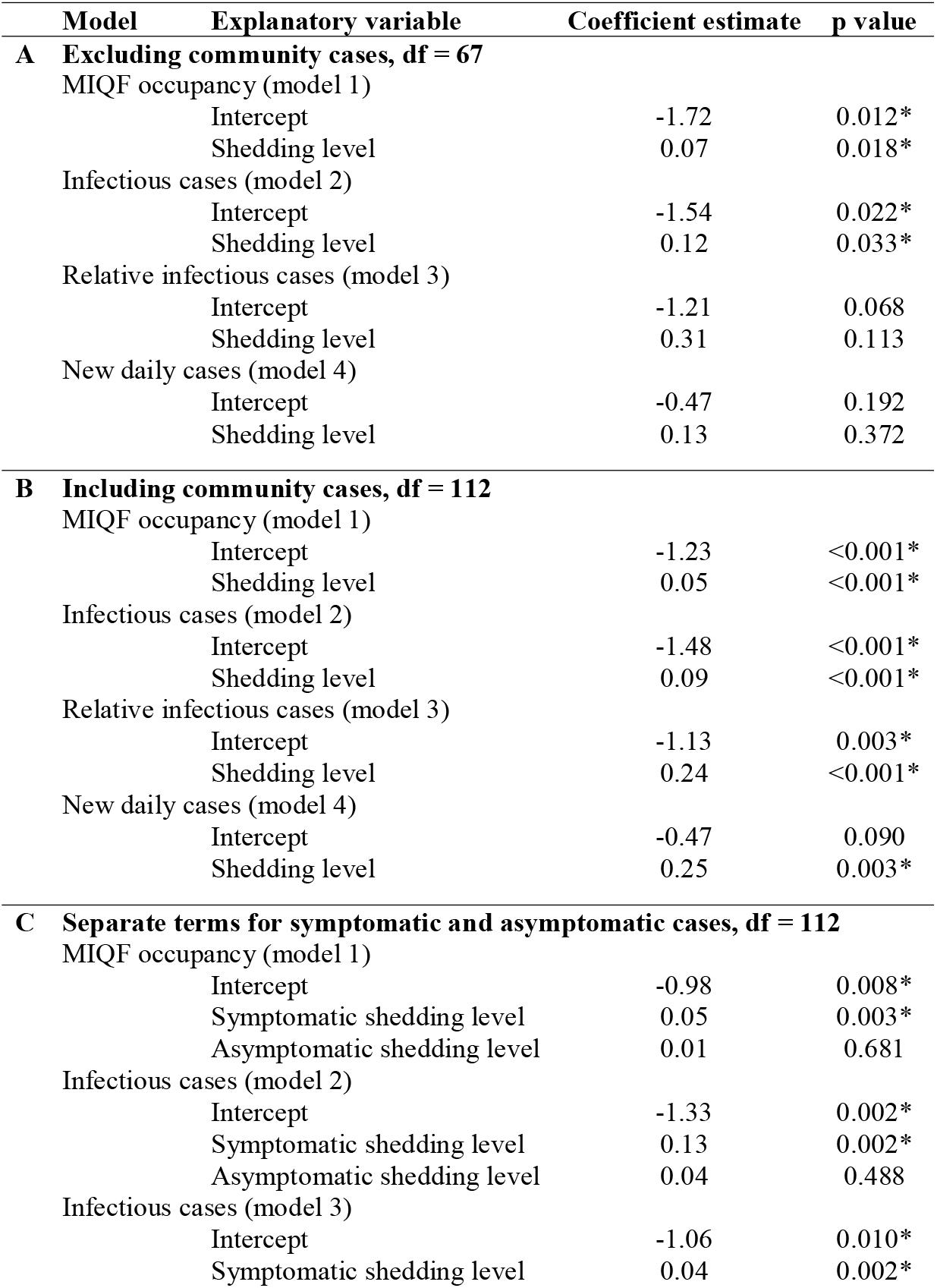
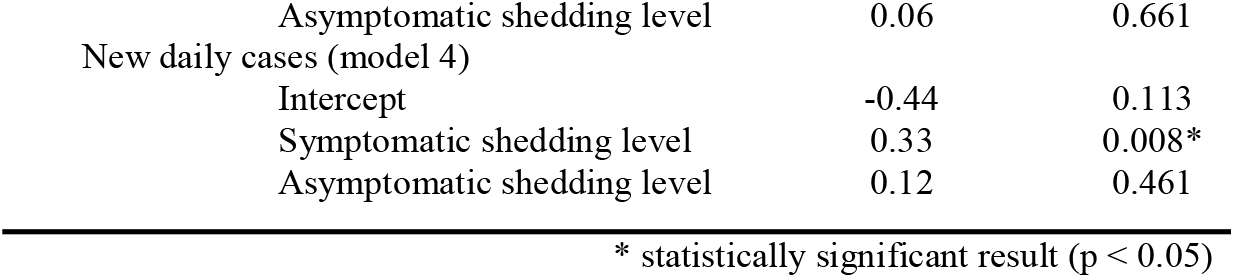
Logistic regression model parameter estimates for the probability of detecting SARS-CoV-2 RNA in wastewater. Table S2A describes the logistic regression models excluding the community cluster period. Table S2B describes the logistic regression models including the community cluster period. Table S2C describes the logistic regression models where symptomatic cases and asymptomatic cases are considered separately. Df is degrees of freedom

**Supplementary Figure S1.**
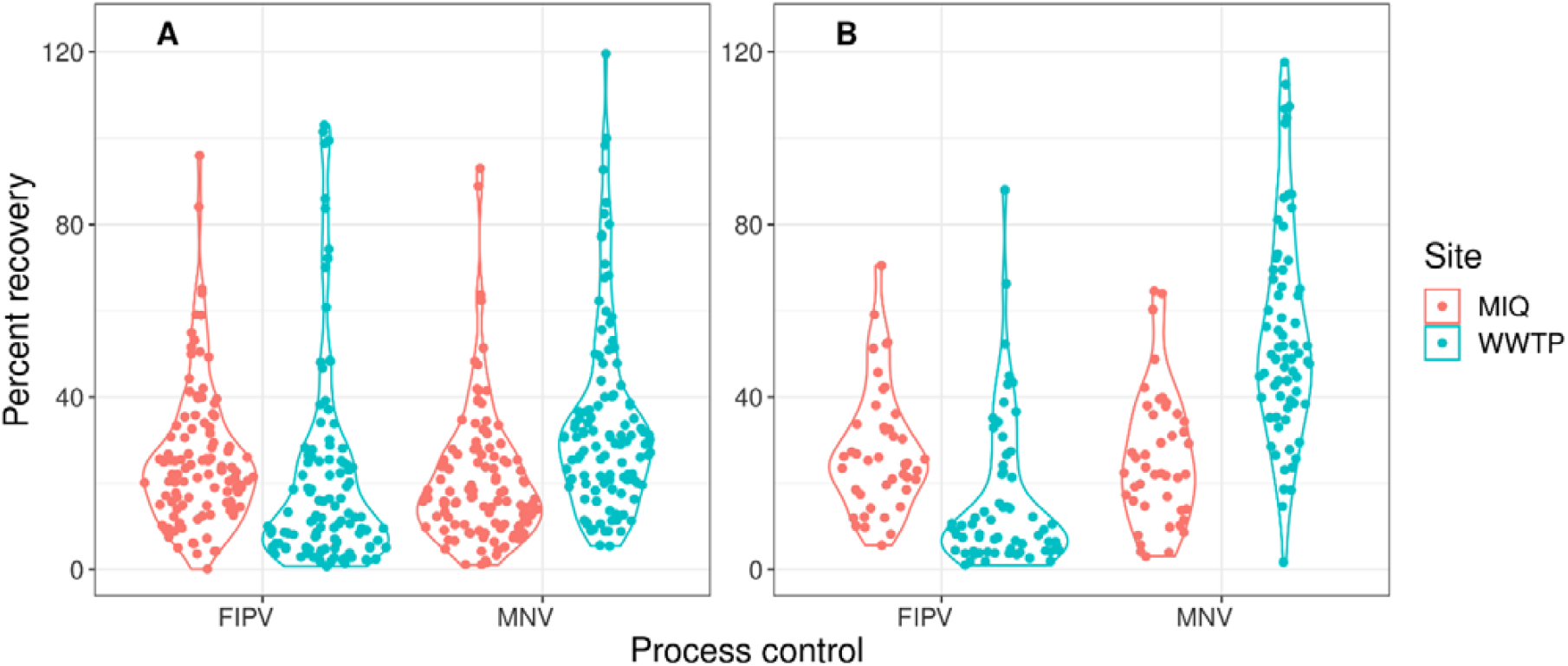
Distribution process control recovery rates. Feline infectious peritonitis virus (FIPV) and murine norovirus (MNV) recovery, from MIQF (red) and WWTP (blue) samples using A) 200 µL extraction volume; B) 50 µL extraction volume is shown. Note that 50 µL extractions were not performed for all samples hence fewer datapoints.

**Supplementary Table S3.** Bootstrap 95% confidence intervals for recovery of the process controls, feline infectious peritonitis virus (FIPV) and murine norovirus (MNV), from MIQF and WWTP samples using 200 µL extraction volume; 50 µL extraction volume. Note that 50 µL extractions were not performed for all samples, hence fewer datapoints. Bootstrap confidence intervals were estimated from 10000 bootstrap samples.

| Volume extracted | Location | Number of samples | Process control | Mean | 95% Confidence interval |  |
| --- | --- | --- | --- | --- | --- | --- |
|  |  |  |  |  | Lower limit | Upper limit |
| 200 μL | MIQF | 113 | FIPV | 25.4% | 22.5 | 28.4 |
|  | MIQF | 113 | MNV | 20.9% | 18.2 | 23.9 |
|  | WWTP | 113 | FIPV | 20.8% | 15.6 | 25.2 |
|  | WWTP | 113 | MNV | 34.0% | 30.0 | 38.3 |
| 50 μL | MIQF | 44 | FIPV | 27.7% | 23.7 | 32.1 |
|  | MIQF | 44 | MNV | 25.8% | 23.1 | 32.3 |
|  | WWTP | 65 | FIPV | 17.7% | 12.9 | 23.5 |
|  | WWTP | 70 | MNV | 54.7% | 49.5 | 61.7 |

